# Aligning staffing schedules with testing and isolation strategies reduces the risk of COVID-19 outbreaks in carceral and other congregate settings: A simulation study

**DOI:** 10.1101/2021.10.22.21265396

**Authors:** Christopher M. Hoover, Nicholas K. Skaff, Seth Blumberg, Rena Fukunaga

## Abstract

COVID-19 outbreaks in congregate settings remain a serious threat to the health of disproportionately affected populations such as people experiencing incarceration or homelessness, the elderly, and essential workers. An individual-based model accounting for individual infectiousness over time, staff work schedules, and testing and isolation schedules was developed to simulate community transmission of SARS-CoV-2 to staff in a congregate facility and subsequent transmission within the facility that could cause an outbreak. Systematic testing strategies in which staff are tested on the first day of their workweek were found to prevent up to 16% more transmission events than testing strategies unrelated to staff schedules. Testing staff at the beginning of their workweek, implementing timely isolation following testing, limiting test turnaround time, and increasing test frequency in high transmission scenarios can supplement additional mitigation measures to aid outbreak prevention in congregate settings.

**Article summary line:** Aligning routine testing with work schedules among staff in carceral facilities and other congregate settings can enhance the early detection and isolation of COVID-19 cases, limiting the potential for staff to inadvertently trigger outbreaks in high-risk settings.

## INTRODUCTION

Throughout the COVID-19 pandemic, outbreaks in congregate settings such as skilled nursing facilities (1), homeless shelters (2–5), and carceral (e.g., prisons and jails) facilities (6) have been devastating. Staff have inadvertently served as a conduit for introducing SARS-CoV-2, the virus that causes COVID-19, from the community to people in congregate settings (6–8). As such, routine testing of staff and subsequent isolation of infectious staff is essential to mitigate case importation among resident populations and staff-to-staff transmission. Prior analyses suggest that routine SARS-CoV-2 screening testing is one approach to reduce transmission in homeless shelters (9), in healthcare settings (10), and during airline travel (11).

In correctional and detention facilities, preventing spillover from the community to facility staff and subsequently into resident populations remains one of many challenges to limit SARS-CoV-2 transmission (12). Having a robust and responsive testing and isolation strategy remains essential to a facility’s success in preventing transmission. As of October 15, 2021 the Centers for Disease Control and Prevention’s (CDC) Interim Public Health Recommendations for Fully Vaccinated People recommends that fully vaccinated people who have come into close contact with someone with suspected or confirmed COVID-19 be tested 5-7 days after exposure and wear a mask in public indoor settings for 14 days, or until they receive a negative test result (13). Due to the high risk of SARS-CoV-2 transmission in congregate settings (6), questions remain around optimal testing policies for staff, regardless of vaccination status, with reports of infections in vaccinated persons in large public gatherings (14), as well as in congregate settings such as health care (15), and correctional (16) facilities.

At this time, the CDC Interim Guidance for SARS-CoV-2 Testing in Correctional and Detention Facilities (17) does not specify when staff should be tested during the workweek to minimize the spread of SARS-CoV-2 via rapid identification and isolation of new staff cases. The timing of systematic testing in relation to work schedules and variable infectiousness profiles could have profound importance for designing optimal systematic testing strategies and for informing downstream activities to prevent transmission, such as rapid identification and isolation of positive staff cases. Testing early in the work week may miss recently acquired infections and lead to staff working around the time of their peak infectiousness. However, testing later in the work week risks missing infectious individuals who are then allowed to work several days prior to being tested and isolated.

This study examines the relationship between work schedules, testing schedules, and within-facility transmission. An analytic framework to estimate the effect of variable testing frequencies and turnaround time between test administration and isolation on SARS-CoV-2 transmission is presented. In addition, an individual-based model which incorporates work and testing schedules influenced by those observed in operations records collected by the California Department of Corrections and Rehabilitation (CDCR) is used to simulate community acquisition of SARS-CoV-2 by staff and subsequent transmission in a congregate setting. Simulations exploring the impact of aligning testing schedules with work schedules are conducted across testing frequency, background community infection rate, and within-facility transmission rate.

## METHODS

### Model framework and parameterization for SARS-CoV-2

Building on previous work investigating the effects of non-pharmaceutical interventions (18) and testing (19) on the transmission of infectious diseases, individual contributions to SARS-CoV-2 transmission through time were modeled from an infectiousness profile, ***β***_***t***_, generated from key biological parameters of the virus that determine the distribution of infectiousness over time. The probability density function of the triangle distribution was used to model ***β***_***t***_, with infectiousness beginning after the latent period, ending after the duration of the infectious period, and peaking at some point in between (***a*** = ***t***_***latent***_, ***b*** = ***t***_***total***_ where ***t***_***total***_ = ***t***_***infections***_ + ***t***_***latent***_, ***c*** = ***t***_***peak***_, and a<c<b; Fig 1a).

**Figure 1.**
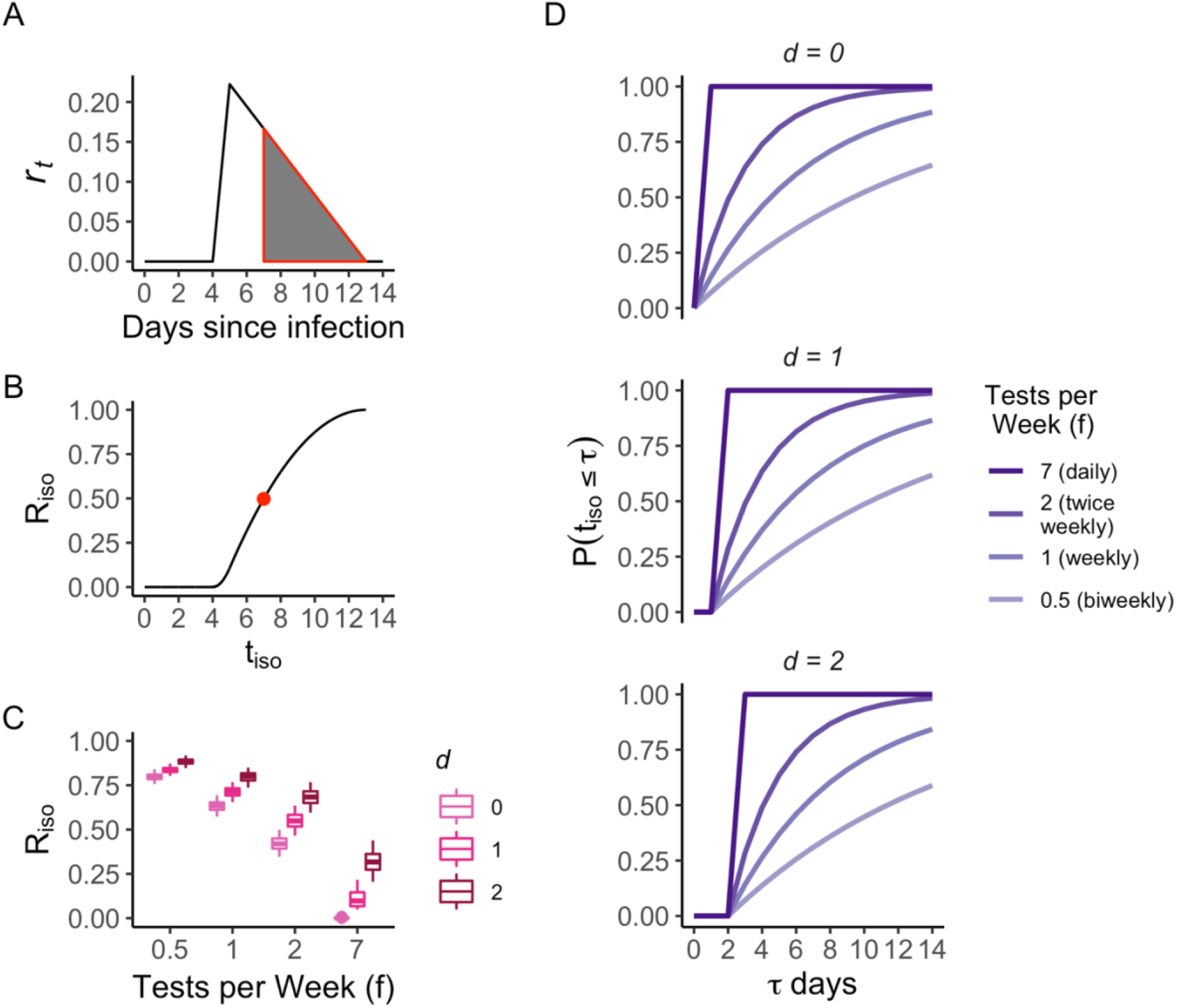
Analytic framework exploring effects of variable infectiousness through time, testing frequencies, and delays on SARS-CoV-2 transmission. A) Example infectiousness profile for ***ℛ*** = ***1, t***_***latent***_ = ***4, t***_***incubation***_ = ***5, t***_***infections***_ = ***9***, with line indicating infectiousness (*r_t_*) through time and shaded area demonstrating infectiousness slice removed if ***t***_***iso***_ = ***7***, leading to ***ℛ***_***iso***_ = **0. 50**. B) ***ℛ***_***iso***_ as a function of ***t***_***iso***_ with same parameters as in A and point indicating scenario depicted in A. C) Boxplots showing distributions of ***ℛ***_***iso***_ as a function of testing frequency, ***f***, and delay in obtaining test results, ***d***, incorporating uncertainty in ***t***_***latent***_, ***t***_***incubation***_, and ***t***_***infections***_ by drawing ***n*** = ***100*** parameter sets for each, with baseline ***ℛ*** = ***1***. Boxplots indicate median, interquartile range, and full range of values of ***ℛ***_***iso***_. D) Probability isolation occurs as a function of testing frequency, ***f***, delay in obtaining test results, ***d***, and days from exposure to isolation *τ*, i.e. ***t***_***iso***_ ≤ ***τ***, demonstrating that delays in obtaining test results substantially reduce the probability of prompt isolation, particularly among most frequent testing scenarios.

The viral dynamics of SARS-CoV-2 make control efforts challenging, as high infectiousness in the absence of symptoms is common (20–22). In terms of the infectiousness profile for SARS-CoV-2, this means that peak infectiousness (***t***_***peak***_) tends to coincide with the onset of symptoms (for cases that are symptomatic), but occurs after completion of the latent period (i.e. ***t***_***peak***_ **≈ *t***_***incubation***_ and ***t***_***incubation***_ > ***t***_***latent***_) (22). The expected number of new cases generated by an individual at time ***t*** is thus ***r***_***t***_ = **ℛ*β***_***t***_, where ***ℛ*** is the effective reproduction number, here defined as the expected number of cases generated in a facility by a new case over the duration of their infectious period, assuming they spent their entire infectious period in the facility. In the absence of other interventions, the model therefore assumes that new cases are most likely to be generated around ***t***_***peak***_ when infectiousness (viral load) is highest. Table 1 lists the distributions of ***t***_***incubation***_, ***t***_***latent***_, and ***t***_***infections***_ used here.

**Table 1:** Distributions and parameter values used in analytic framework and model simulations. The latent period is defined as the time between exposure and onset of infectiousness, the incubation period as the time between exposure and both symptoms and peak infectiousness (even in the absence of symptoms), and the infectious period as the total time a case is infectious.

| Parameter | Distribution | Source |
| --- | --- | --- |
| Incubation Period ($t_{incubation}$) | Log normal (1.63, 0.5) | (23) |
| Latent Period ($t_{latent}$) | $t_{incubation} - \text{Uniform}(0, 2)$ | (22,24) |
| Infectious Period ($t_{infectious}$) | Uniform (7, 10) | (22,24) |

In the presence of interventions that isolate infectious individuals prior to ***t***_***total***_, for example through contact tracing, self-isolation following the onset of symptoms, or isolation following a positive test result, the effect of isolation on ***ℛ*** can be directly estimated from the time to isolation as 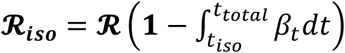, where ***t***_***iso***_ is the time at which isolation occurs. Reducing ***ℛ***_***iso***_ via improved contact tracing or more frequent testing can thus be represented as removing a larger slice from the overall infectiousness triangle by reducing ***t***_***iso***_ (Fig 1a).

Figure 1b shows the relationship between ***ℛ***_***iso***_ and ***t***_***iso***_ is sigmoidal, implying earlier isolation is incrementally more effective and the benefits of isolation level off later in the infectious period. Other interventions that reduce ***ℛ*** across all levels of infectiousness such as levels of vaccination coverage, wearing a mask, or reducing the contact rate between infectious and susceptible individuals can also be accommodated simply by multiplying ***ℛ*** by a constant.

The test frequency, ***f***, is defined as the average number of tests per week. Assuming testing is done randomly through time and is independent of symptoms or known contacts, the probability of going ***t*** days without being tested can be estimated as (**1** − ***f***/***7***)^***t***^, where, for example ***f*** = **1** if testing is conducted weekly. The probability that isolation has occurred by day ***τ*** after onset of infectiousness can then be estimated as ***P***(***t***_***iso***_ ≤ ***τ***) = **1** − (**1** − ***f***/***7***)^***τ***^ if isolation occurs immediately after testing. Given substantial turnaround times between testing and isolation, particularly when relying on nucleic acid amplification tests (NAATs), the delay, ***d***, between testing and isolation can also be incorporated as: ***P***(***t***_***iso***_ = ***τ***) = **0 for *τ*** < ***d*** and ***P***(***t***_***iso***_ = ***τ***) = **1** − (**1** − ***f***/***7***)^***τ***−***d***^ ***for τ*** ≥ ***d***. Figure 1d shows that such delays have a detrimental effect on the probability of achieving prompt isolation, particularly by making isolation prior to the delay (***t***_***iso***_ < ***d***) impossible.

Testing frequency and delays obtaining test results can also be incorporated into estimation of **ℛ**_***iso***_, with the reduction in ***ℛ*** due to isolation estimated from infectiousness on day ***t*** weighted by the probability of being isolated on day ***t***. Discretizing, this gives:

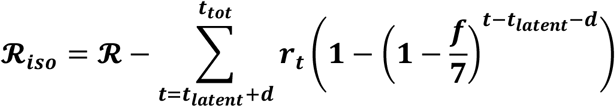

Figure 1c shows distributions of ***ℛ***_***iso***_ derived from 100 random draws sampling from uncertainty in the SARS-CoV-2 latent, incubation, and total infectious periods, across test frequencies ranging from daily (***f*** = ***7***) to biweekly (***f*** = **0. 5**) and delays in obtaining test results from 0 to 2 days. *ℛ*_*iso*_ is similar when testing every day (*f* = 7) with a two-day turnaround time for test results (*d* = 2) vs testing twice per week (*f* = 2) with immediate test results (*d* = 0) (Fig 1c, median *ℛ*_*iso*_ (*d* = 0, *f* = 2) = 0.42 and *ℛ*_*iso*_ (*d* = 2, *f* = 7) = 0.33, respectively), again reiterating the importance of reducing delays in obtaining test results.

### Individual-based model simulations

#### Model setup

The framework described above demonstrates the benefits of high test frequency and limited delays between testing and isolation to prevent SARS-CoV-2 transmission, but it is not capable of investigating how testing and staffing schedules should be configured to optimally prevent transmission in a congregate facility. An individual-based model building on this framework and incorporating staff working and testing schedules was therefore developed to simulate SARS-CoV-2 transmission within a congregate facility. In a modeled facility, ***w*** staff are assigned a work schedule that determines time frames when they are in the facility interacting with residents and other staff working at the same time. The function ***𝒲***(*w*_*it*_) is defined as an indicator function for whether staff member ***i*** is working at the facility at time step ***t***. In addition to their work schedule, all staff are assigned a testing schedule, encoded by function ***τ***(*w*_*it*_), with different testing schedules discussed further below. The model is simulated for 180 days with three 8-hour time steps per day (*t*_*sim*_ = 540) with ***w*** = 700 staff, with each time step corresponding to a work shift as described below.

Staff move through susceptible (S), exposed (E), infected (I), and recovered (R) states, with the infected state corresponding to time when ***β***_***it***_ > ***0***. Recovered staff are assumed to remain in state R and not return to state S due to the relatively short time frame of the simulation. Parameters for newly exposed staff are drawn to determine ***t***_***latent***_, ***t***_***incubation***_, and ***t***_***infections***_, from which an infectiousness profile, ***β***_***it***_ is generated. Tested staff produce a positive test result if ***β***_***it***_ > ***0*** and ***τ***(*w*_*it*_) = **1**, at which time they enter an isolated (O) state immediately if ***d*** = **0**. If there is a delay between test administration and the test result (***d*** > **0**), staff first enter a tested (T) state before O, during which time they may continue to work while infectious, inadvertently exposing others in the facility. Staff in state O are restricted from working for 10 days (***𝒲***(*w_it_*) = **0** for 10 days) and are not required to undergo systematic testing for 90 days following a positive result (***τ***(*w*_*it*_) = ***0*** for 90 days).

Assuming constant ***ℛ*** across all individuals and through the duration of the simulation, the expected number of infections in the facility at time step ***t*** caused by individual ***i*** is ***r***_***it***_ = ***ℛ β_it_𝒲***(***w***_***it***_). Three separate values of ***ℛ*** (0.5, 1.0, 1.5) were simulated to explore different levels of containment and effectiveness of mitigation strategies within facilities. Staff may acquire infection from the community according to the community prevalence when they are not working (***𝒲***(***w***_***it***_) = 0) or from fellow staff while working (***𝒲***(***w***_***it***_) = 1) where the force of infection is 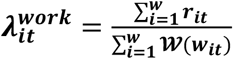. The expected number of infections in the facility generated by staff is estimated from each simulation as: 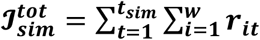.

### Staffing and testing strategies

CDCR collects operations records for custody staff including information on workdays (e.g., Mon-Thurs), work shifts (e.g., morning, evening, night), and SARS-CoV-2 testing schedules. We used this information to generate a realistic representation of staff working schedules in model simulations by sampling from standard work schedules identified among custody staff using K-means clustering.

Two experimental testing strategies were considered in model simulations. Under a random testing strategy, testing for each worker occurs at random during their work shifts depending on the frequency (i.e. with *f* = 2, workers would be tested during two of their shifts, chosen at random each workweek). Under a systematic testing strategy, each worker is always tested on the same day(s) of their shift each week. For *f* = 1, systematic testing always occurs on the first day of their workweek; for *f* = 2, systematic testing always occurs on the first and third days; and for *f* = 4, testing occurs on each of the first four workdays in a workweek.

All tests conducted when ***β***_*it*_ > 0 are assumed to return a positive result (25–27) and no testing other than systematic screening testing occurs. The total number of tests conducted in each simulation is recorded as: 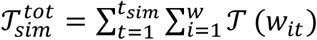 Combined with the expected number of cases in the simulation, the incremental test effectiveness ratio (ITER) is estimated as: 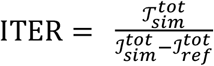, where 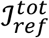 is the number of infections in a reference scenario with no testing. The ITER can be interpreted as the number of tests needed to prevent one infection in the simulation scenario being evaluated.

All simulations, analyses, and visualizations were compiled in R software version 4.0.4 (28) with aid from the tidyverse (29), triangle (30), and patchwork (31) packages. Code is available at https://github.com/cmhoove14/Congregate-Staff-Testing.

## RESULTS

### Staff working and testing schedules

Four typical staff workweek schedules were identified using K-means clustering from CDCR operations records. Most common was a four-day workweek in which the staff member worked four consecutive days (e.g., Monday-Thursday) and a variable fifth day, though the first day of the workweek varied across staff (Figure 2). Work shifts also tended to show consistent patterns. Staff typically worked eight hours during either the morning, evening, or night shift, though alternating between morning and evening shifts, and taking on an additional shift was also common. These work schedules were used to generate a realistic representation of staff schedules in model simulations. Tests were most often administered on Tuesdays (if the staff had Tuesday in their typical workweek) regardless of whether it was the first day of the staff’s workweek. Testing on Wednesday and Thursday was also common across work schedules. Test results were usually returned on the same day or the day after specimen collection and almost all test results were received within 2 days of specimen collection.

**Figure 2.**
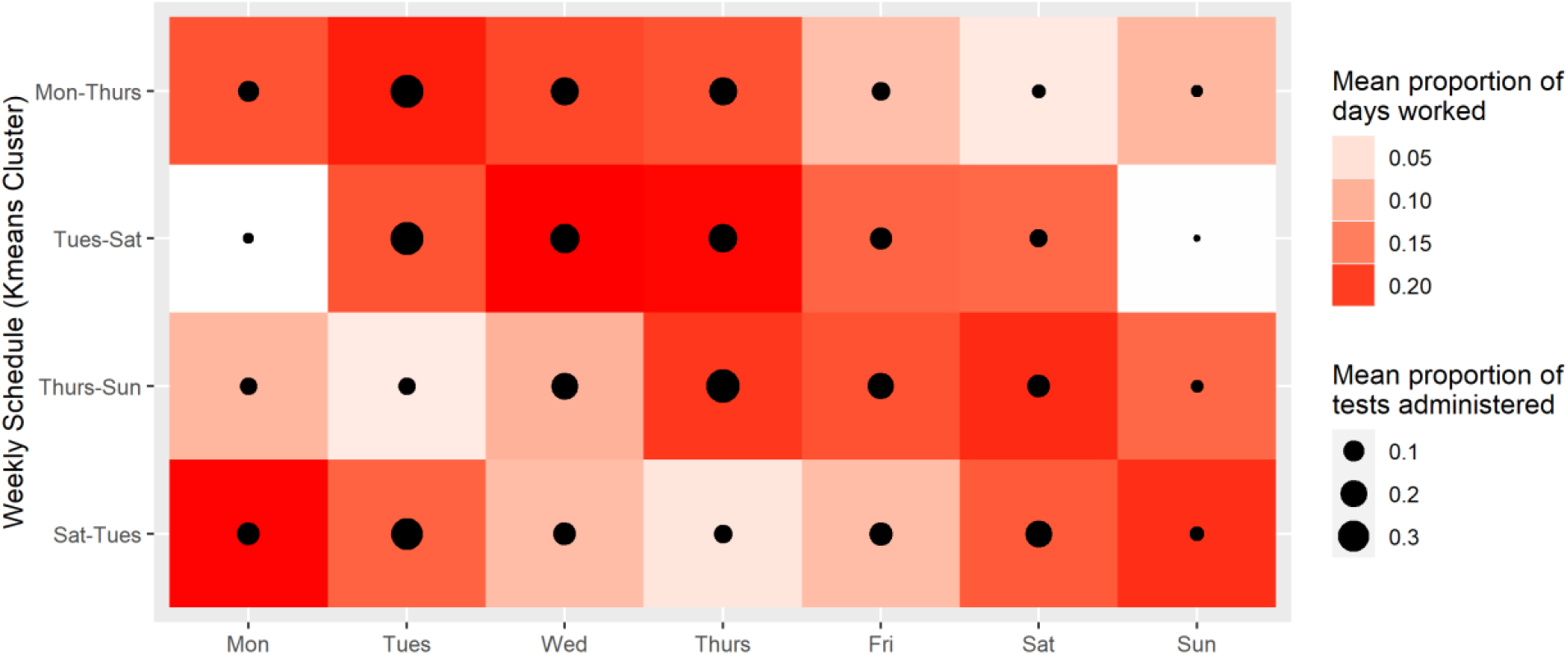
Staff work and testing schedules. Four typical weekly work schedules (y-axis) were identified among CDCR custody staff. These include a Monday to Thursday workweek (21% of staff), a Tuesday to Saturday workweek (33% of staff), a Thursday to Sunday workweek (22% of staff), and a Saturday to Tuesday workweek (24% of staff). The red shading shows the mean proportion of staff workdays that consist of a particular day of the week (x-axis; i.e. darker shades of red indicate that staff with the specified schedule more commonly worked on that day). The size of the black circles represents the mean proportion of the total number of tests administered to each group that were given on the specified day.

### Simulation Results

Systematic testing strategies were found to consistently outperform random testing strategies in terms of preventing infections within simulated facilities. Figure 3 shows a comparison of the number of infections 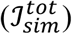 when implementing random vs. systematic testing strategies across testing frequencies, levels of community prevalence, and within-facility ***ℛ*** with either no delay or a one-day delay between test administration and isolation of infectious workers. In the highest transmission scenario (*CP* = 1%, ***ℛ*** = 1.5), no testing led to a median 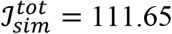 (IQR 108.13 - 114.73) expected transmissions. Testing randomly once per week with no delay to isolation resulted in a median 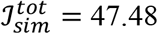 (IQR 44.52 - 50.3; Fig 3 right panel, rightmost yellow circle), whereas testing systematically on the first day of the work week with no delay to isolation resulted in 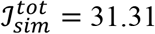 (IQR 29.48 - 33.21; Fig 3 right panel, rightmost yellow square). However, systematic testing that is accompanied by a one day delay leads to 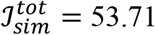 (IQR 50.98 - 55.87; Fig 3 right panel, rightmost yellow cross).

**Figure 3.**
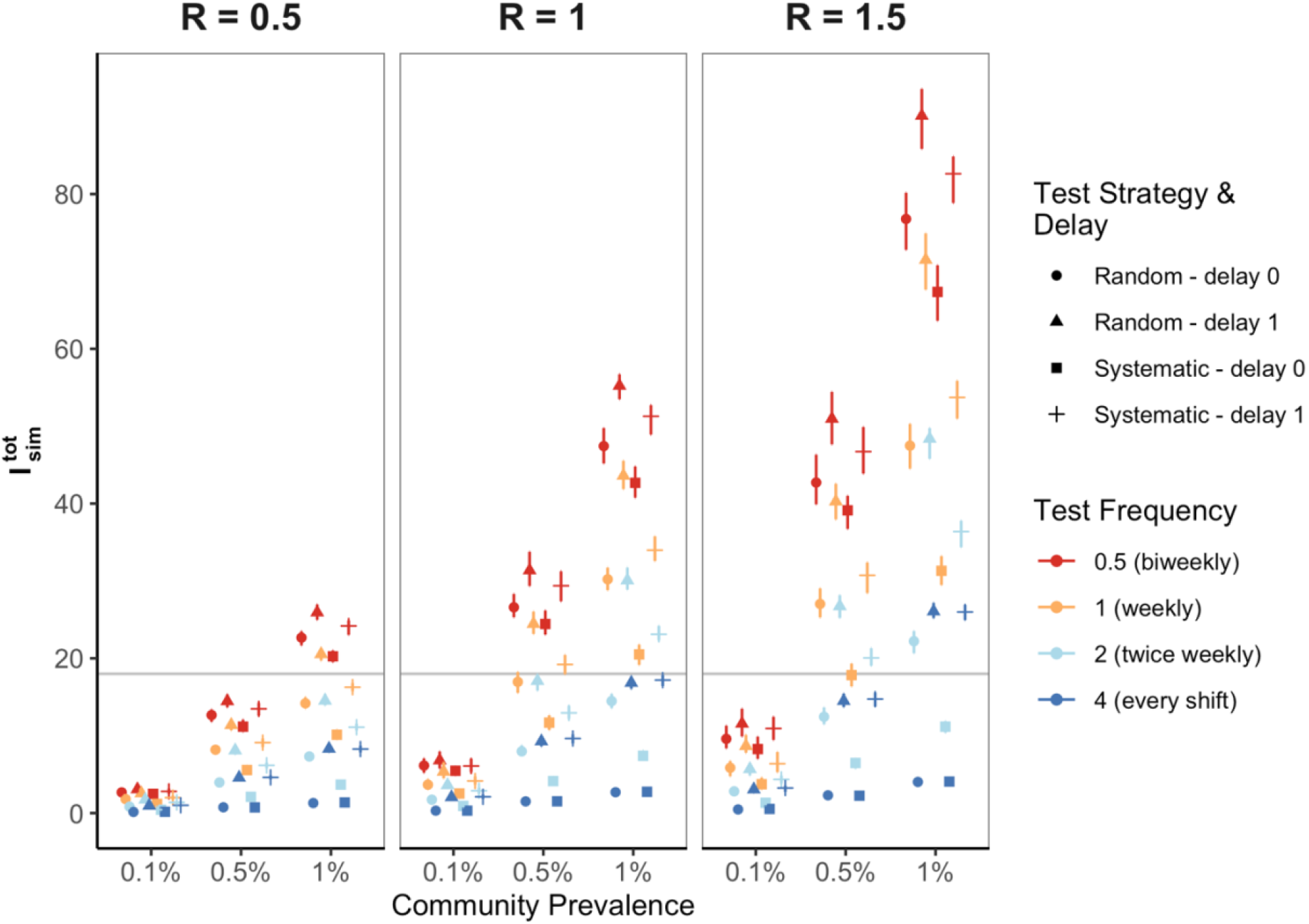
Number of expected infections generated in a facility from model simulations comparing random and systematic testing strategies across transmission scenarios, test frequencies, and delays isolating infectious individuals who have tested positive. Systematic testing strategies (◼, ┼) prevent more infections than random strategies (•, ▴) across all transmission scenarios (indicated by community prevalence across the x axis and by reproduction number across the panels) and test frequencies (indicated by different colored symbols with blue corresponding to the highest test frequency of 4 tests per week and red the lowest test frequency of biweekly testing). More transmission events are expected in transmission scenarios with higher within-facility ***ℛ*** and higher community prevalence. Preventing delays between testing and isolation of positives (squares compared to crosses and triangles compared to circles) and increasing test frequency (red=lowest frequency, blue=highest frequency) also reduces the number of transmission events. The horizontal gray line serves as a reference to assess the testing frequency needed to maintain 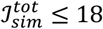 (corresponding to one transmission event every ten days) across different transmission scenarios. Error bars represent the interquartile range of 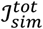 derived from 100 simulations per scenario run for 180 days among 700 staff.

Across all transmission scenarios, biweekly systematic testing with no delay to isolation averted an average of 40% of transmissions that would have occurred with no testing, while random testing averted an average of 33% of transmissions. For weekly frequency, systematic testing averted an average of 71% of transmissions versus 57% of transmissions when testing randomly; and for twice weekly testing, systematic testing averted and average of 90% of transmissions versus 80% of transmissions when testing randomly.

The horizontal gray line in Figure 3 demonstrates a potential threshold number of infections to avoid exceeding at 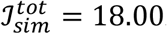. This threshold corresponds to an average of one transmission event within the simulated facility every ten days. Implementing a systematic– rather than random–testing strategy can be sufficient to prevent 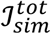 from exceeding such a threshold without changing the frequency in many transmission scenarios (e.g. compare circles to squares of the same color in Figure 3) though in the highest transmission scenarios, greater than twice-weekly testing may be needed. Table 2 additionally shows the testing frequency in tests per week under a systematic testing strategy necessary to ensure that the upper quartile of expected transmission events is maintained below this threshold.

**Table 2:**
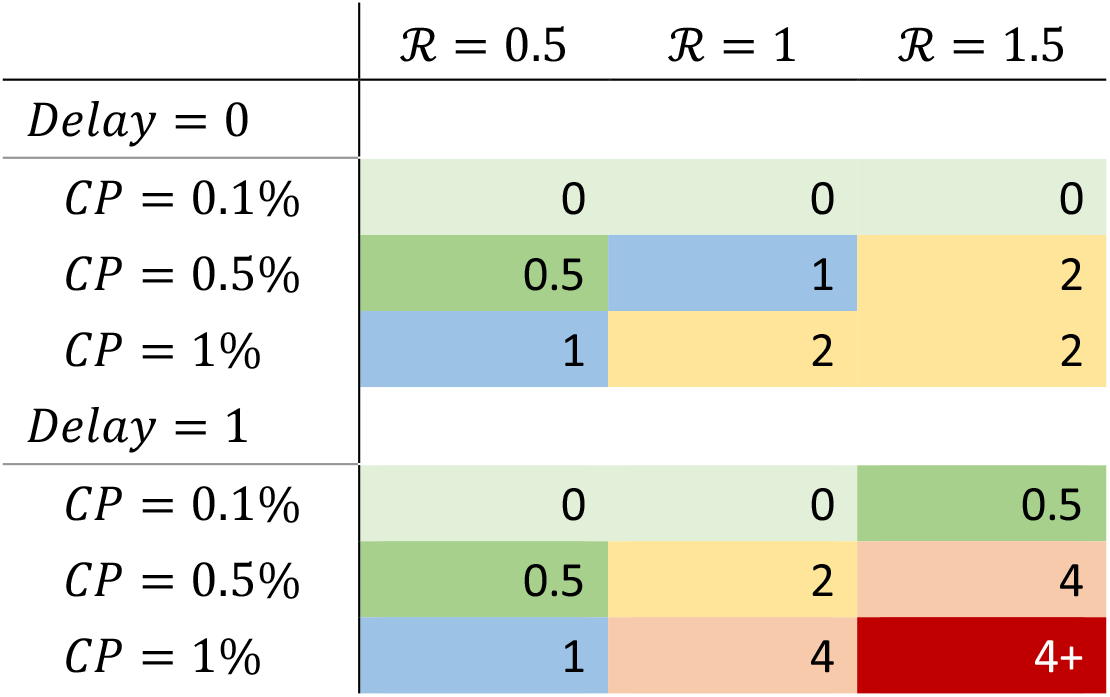
Test frequency (tests per week) under a systematic testing strategy needed to maintain the upper quartile of expected infections in the simulated facility below a threshold of 1 every ten days across transmission scenarios conveyed by the within-facility basic reproduction number (***ℛ***), community prevalence (CP), and delays between testing and isolation of infectious workers

An alternative threshold approach to aid decision-making, particularly in resource-constrained settings, is the ITER, interpreted as the number of tests needed to prevent an infection in the simulated facility. Figure 4 shows estimates of the ITER across transmission scenarios, test strategies, and test frequencies. In the highest transmission scenario (***ℛ*** = 1.5, 1% community prevalence), testing systematically on the first day of every other work week with no delay (*f* = 0.5, *d* = 0, Fig 4, see squares) leads to ITER = 180.89 (IQR 168.01 - 196.5), while increasing test frequency to weekly (f = 1) results in ITER = 181.72 (IQR 178.78 - 186.08), to twice weekly (f = 2): ITER = 293.02 (IQR 288.91 - 295.6), and to every shift (f = 4): ITER = 545.36 (IQR 541.58 - 550.61). These values approximately correspond to test positivity rates of 0.55%, 0.55%, 0.34%, and 0.18% due to the interpretation of the ITER as the number of tests per positive result. Figure 4 also provides an example reference line at *ITER* = 400, corresponding to an approximate 0.25% test positivity, to demonstrate how testing frequency may be determined from the transmission scenario and target ITER, which may be influenced by the number of tests available.

**Figure 4.**
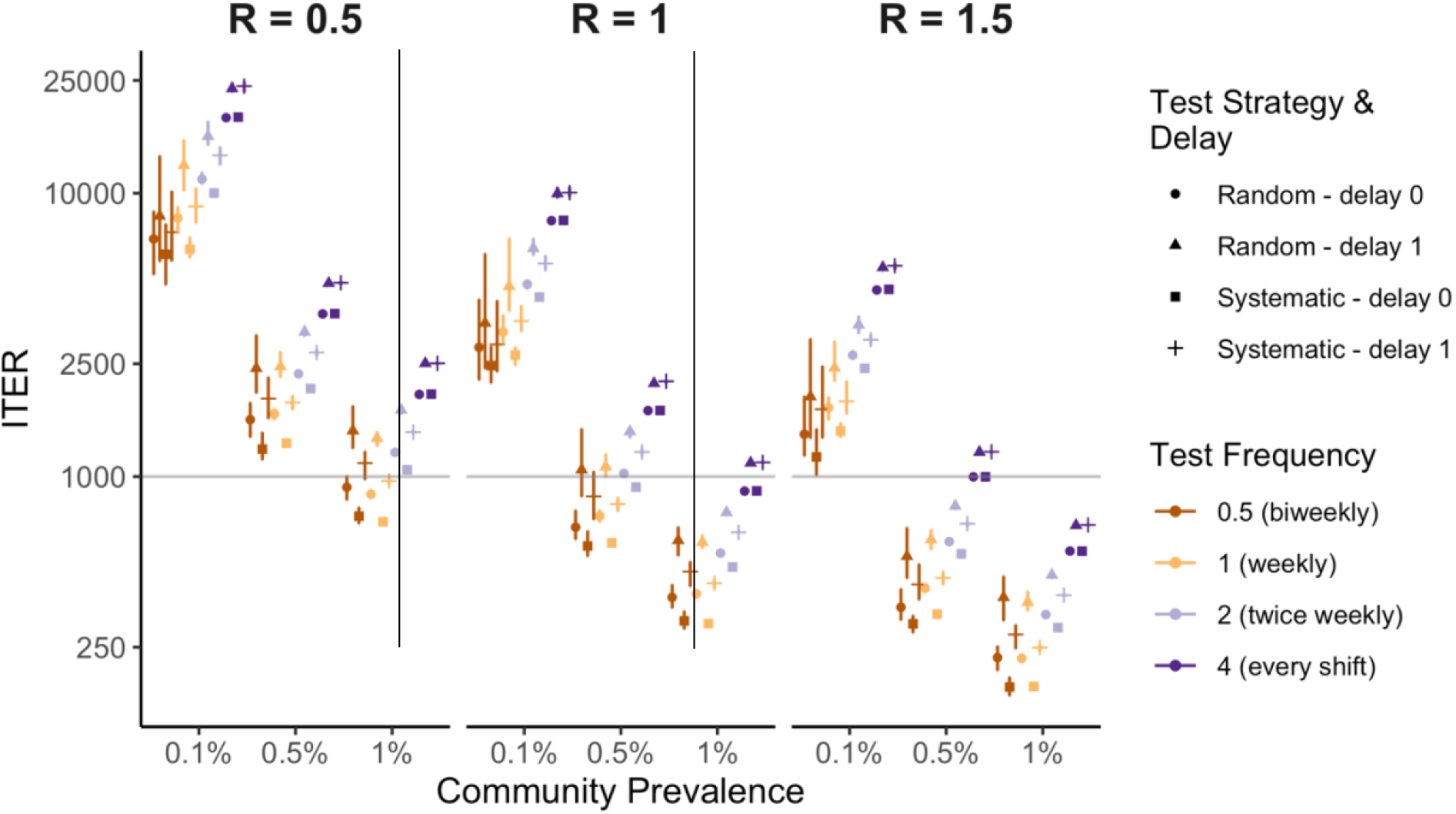
Incremental test effectiveness ratio (ITER) from simulations across transmission scenarios and testing frequencies and strategies. The ITER remains relatively low in higher transmission scenarios even at high (***f*** = ***4***) testing frequencies, potentially favoring such high-frequency testing strategies when within-facility transmission (***ℛ***) and/or community prevalence are high. The y-axis is log-transformed and the horizontal line at ***ITER*** = ***1000*** is provided to aid visual comparison across scenarios. Error bars represent the interquartile range of expected infections derived from 100 simulations per scenario.

## Discussion

This study builds on previous modeling and simulation analyses to demonstrate that systematic testing strategies with limited delays between test administration and isolation of infectious individuals can limit SARS-CoV2 transmission. Building on this, testing schedules that are aligned with working schedules are found to prevent more transmission events than random testing strategies or those with a delay between testing and isolation. A major benefit of such strategies is that they do not require higher testing frequency, only a change in timing of when testing occurs. As such, there may be substantial value in implementing systematic rapid testing at the beginning of the work week for staff working in facilities at high risk for SARS-CoV-2 transmission such as carceral facilities, skilled nursing facilities, and homeless shelters.

For SARS-CoV-2, the occurrence of pre- and asymptomatic transmission calls for systematic testing to be a key component of prevention strategies. Confirmatory testing with sensitive NAATs may be necessary in scenarios where less sensitive rapid antigen tests with quicker turnaround time are used as an initial screen. Additionally, increasing the frequency of testing may be necessary in settings with high community prevalence or the opportunity for rapid spread of the virus within a facility (e.g. highly transmissible variants, low vaccination rates, inadequate mitigation practices). Lower thresholds than one expected infection event per ten days may also be necessary to prevent outbreaks in carceral facilities and other congregate settings. A prior analysis of publicly available CDCR case data estimated 46% of 118 SARS-CoV-2 introductions into resident populations from April 2020 to March 2021 across 35 facilities resulted in outbreaks of greater than 10 resident cases (32), though this estimate includes data from early in the pandemic when there were more fully susceptible individuals, fewer protocols to reduce transmission, limited testing resources, and lower vaccination coverage.

This study also utilized the ITER as a per-test measure of effectiveness for systematic testing across a range of frequencies and transmission scenarios. In resource-constrained environments in which tests are difficult to acquire (e.g., limited supply/funds), the ITER and its relationship to test positivity may be used to guide decisions on test frequency. The ITER may also be useful in situations where further data on the cost per COVID-19 case and cost per test conducted are available. In this case, the product of the ITER and the cost per test conducted provides the cost per case avoided due to the testing program. For facility management, any testing program that results in a lower cost per case avoided than cost per COVID-19 case would likely be deemed cost effective.

Even though systematic testing strategies reduce within-facility transmission, they are not capable of preventing all transmission events. Systematic testing represents one tool of many that could be considered for implementation to prevent SARS-CoV-2 infections in congregate facilities. Facility-wide vaccination, universal masking, rapid isolation of COVID-19 cases, quarantine of individuals after a potential exposure, avoiding crowds, physical distancing, and proper ventilation, all play an important role in mitigating SARS-CoV-2 transmission in carceral facilities and other congregate settings (33). However, sometimes low vaccine acceptance rates among both residents and staff in correctional settings coupled with more transmissible SARS-CoV-2 variants puts this population at continued risk of localized outbreaks. Implementing routine, systematic testing of staff for early identification of COVID-19 cases (including infections in vaccinated persons) is another layer of intervention that can prevent outbreaks from occurring within congregate facilities.

There are several notable limitations to this model. First, staff are not the only source of infection, as there are other potential sources of importation into the facility including: intake of new residents, visitation, facility movement, and work programs where residents leave the facility during the day. Second, the exclusion of notable COVID-19 prevention strategies (e.g. universal masking, physical distancing, proper ventilation) and of additional testing due to symptoms or known contacts is a limitation of our model. However, if additional control interventions were implemented, we expect qualitative trends in the expected number of transmission events to persist between testing strategies and frequencies across different transmission scenarios. Third, we do not distinguish between staff-to-staff and staff-to-resident transmission events within a simulated facility, but rather record the total number of transmission events assuming ***ℛ*** remains constant rather than decreasing due to susceptible depletion.

Estimation of staff-staff and staff-resident contact rates or reproduction numbers would enable more precise accounting and simulation of importation events and subsequent transmission within a facility. Fourth, we assume that the probability density function of the triangle distribution is an accurate representation of SARS-CoV-2 viral dynamics and therefore infectiousness through time. Though this function captures the general viral dynamics profile seen previously (19,22), other distributions or functions may also be applicable, though other analyses using more complex infectiousness profiles have yielded similar results (34). Finally, we assume that the community force of infection among staff is constant through time and across individuals. In reality, community prevalence can increase rapidly, necessitating a corresponding increase in test frequency. Furthermore, some staff may be more or less likely to acquire infection in the community or in the facility based on vaccination coverage, compliance with physical distancing and masking policies, their household structure and/or health status, and other behavioral factors.

The modeling and simulation framework presented here is applicable beyond COVID-19 in congregate settings in which outbreaks may be due to community importation of a pathogen. Other applicable settings may include the introduction of hospital acquired infections from newly admitted patients or from hospital staff (35), introduction of other respiratory pathogens such as influenza or pertussis into congregate settings (36), or tuberculosis transmission between communities and populations experiencing incarceration (37). Accurate parameterization of key natural history traits of the pathogen in question such as the latent, incubation, and infectious periods is essential to estimate the impact of nonpharmaceutical interventions such as systematic testing (18). Pathogens other than SARS-CoV-2 that cause symptoms prior to infectiousness (*t*_*incubation*_ < *t*_*latent*_), for instance, may be more effectively controlled at lower cost via symptom screening and subsequent isolation (18).

In conclusion, these results suggest that aligning the timing of testing with regular working schedules for staff in congregate settings, in addition to timely implementation of prevention strategies (e.g., isolation) can improve the efficacy of systematic screening testing. Two metrics, the number of expected within-facility transmission events and the ITER, derived from simulated facilities are presented to inform decisions on the frequency of systematic testing needed in different transmission scenarios to limit transmission under key thresholds. Based on these findings, congregate settings such as carceral facilities, nursing homes, schools, and more may be able to avoid potential outbreaks through systematic testing of staff and other facility residents that is aligned with work schedules and is continued until community transmission or within-facility transmission potential are sufficiently reduced.

## Data Availability

All data produced in the present work are contained in the manuscript and online at https://github.com/cmhoove14/Congregate-Staff-Testing

## Acknowledgements

We acknowledge the California Department of Corrections and Rehabilitation, California Correctional Health Care Services, and the CDC’s COVID-19 Response for supporting this study.

## Disclaimer

The findings and conclusions in this report are those of the author(s) and do not necessarily represent the official position of the U.S. Department of Health and Human Services, the Centers for Disease Control and Prevention, or the authors’ affiliated institutions.

## Biographical sketch

Dr. Hoover is a postdoctoral scholar at the Francis I. Proctor foundation at the University of California, San Francisco. He is interested in using quantitative methods to inform intervention strategies to reduce the global burden of infectious diseases.

